# Hypertension incidence among individuals successfully treated for pulmonary tuberculosis

**DOI:** 10.1101/2024.06.24.24309432

**Authors:** Victoria C Ontiveros, Maia Kipiani, Teona Avaliani, Sara C Auld, Mariam Gujabidze, Russell R Kempker, Hardy Kornfeld, Cassandra Bryan, Moises A Huaman, Matthew J Magee, Argita D Salindri

## Abstract

We reported a cumulative incidence of hypertension of 28% among individuals successfully treated for pulmonary tuberculosis (TB) with no prior hypertension indication at TB treatment initiation (i.e., prevalent hypertension). The cumulative incidence of hypertension among those with ≥1 cavitary lesion was twice the cumulative incidence among those without cavitary lesions.

## INTRODUCTION

Emerging evidence suggests that tuberculosis (TB) disease increases the risk of cardiovascular events during and after TB treatment.^1^ A systematic review conducted by Romanowski et al. also suggested that cardiovascular disease (CVD) is the most frequent cause of long-term mortality in people treated for TB.^2^ Furthermore, cross-sectional studies have shown that people with TB have a high prevalence of risk factors for CVD among people with active TB.^3,4^ However, prospective studies assessing the CVD risk trajectories among people after they have completed TB treatment are scarce as it may require a long follow-up period.

Hypertension, a key mediator in CVD pathophysiology, has been shown to increase the risk of mortality in individuals treated for TB.^5^ A systematic review found that up to an estimated 38.2% of people with active TB have hypertension.^6^ However, most epidemiologic studies to date are cross-sectional or examine the impact on hypertension on TB outcomes while the impact of TB disease on incident hypertension post-TB treatment completion is not well described. Furthermore, little is known which TB-related factors could increase the risk of incident hypertension.

As 11% of patients with active TB are estimated to have a cardiovascular disease^7^ and post-TB mortality rates are nearly three times that of the general population,^2^ there have been calls to understand the directionality of the relationship between active TB and hypertension^6^ and to integrate TB and cardiovascular health care.^8–10^ Targeting non-communicable disease (NCD) prevention efforts like hypertension control may be critical to reducing the chronic impacts of TB.

In this study, we aimed to estimate the prevalence of hypertension at TB treatment initiation and the cumulative incidence of hypertension during and within 12 months post-treatment completion. We also estimated the relationship between TB disease severity and incident hypertension.

## METHODS

### Study Design, Setting, and Participants

We conducted a prospective cohort study among adults successfully treated (i.e., those who received microbial cure or completed treatment) for pulmonary TB at the National Center for Tuberculosis and Lung Diseases in Tbilisi, Georgia, from 2020 to 2022. Eligible participants included adults (≥16 years) with laboratory confirmed (by smear, culture, or rapid molecular test [Xpert]) pulmonary TB. Individuals with a history of lung cancer, prior pulmonary TB treatment, HIV co-infection, and pregnant women were excluded. Participants were recruited at the time of TB treatment completion and followed prospectively at 6- and 12-months post-treatment.

### Study Measures

Our primary outcome of interest was hypertension and data were available from 4 time points: TB treatment initiation (abstracted from medical charts), treatment completion, and at 6- and 12-months post-treatment completion. Prevalent hypertension was defined as 1) a single reading of either systolic blood pressure (SBP) ≥130mmHg or diastolic blood pressure (DBP) ≥80mmHg; or 2) a self-reported history of hypertension diagnosis by healthcare providers at TB treatment initiation. Among those without prevalent hypertension, incident hypertension was defined as ≥2 readings of either SBP ≥130mmHg, DBP ≥80mmHg, or a self-reported hypertension diagnosis at separate follow-up study visits (end of TB treatment, 6- and 12-months post-treatment). Participants were required to have ≥2 blood pressure measurements during follow-up to be considered eligible in hypertension incidence analyses. A sensitivity analysis was performed to evaluate whether measures of association changed substantially when using the WHO-defined hypertension cut-off (i.e., ≥140/90mmHg). To describe changes in SBP, we subtracted SBP absolute values measured at the 12 months post-TB treatment from SBP values measured at treatment initiation.

Indicators of TB severity were determined by chest computed tomography (CT) findings (i.e., presence of cavitary lesions, ≥1 pulmonary lobes with severe [>75%] involvement, and high total lung severity score [i.e., ≥10]). Other covariates included age, gender, drug-susceptibility profile (i.e., drug-susceptible and isoniazid monoresistant TB [DS/Hr TB] vs. rifampicin-resistant and multidrug/extensively drug-resistant TB [RR/MDR TB+]), body mass index (BMI) and BMI changes during TB treatment (BMI increase ≥5% vs. less), glycated hemoglobin (HbA1c), visceral adipose index (VAI)^11^, low-density lipoprotein (LDL), high-density lipoprotein (HDL), and triglycerides. BMI was categorized as underweight (BMI <18.5 kg/m^2^), normal (BMI 18.5–24.9 kg/m^2^), and overweight/obese (BMI ≥25 kg/m^2^).^12^

### Statistical Analyses

We used chi-square or Fisher’s exact tests to assess unadjusted associations between participants’ characteristics and hypertension incidence (p-values were not shown). We used robust Poisson regression models^13^ to estimate cumulative incidence ratios (CIR) of incident hypertension, comparing individuals with ≥1 cavitary lesion vs. no lesions. Covariates included in the final model were based on bivariate association, confounders identified in published literature, and directed acyclic graph. A final multivariable model was determined by selecting a model with the lowest Akaike Information Criterion (Table S1). Mann-Whitney U tests were used to compare median changes of SBP among individuals with hypertension (n=26). We characterized these changes in boxplots produced by using *ggplot2* package.^14^ All analyses were perfomed in R (v.4.2.2; R Core Team 2023)^15^ with p-values <0.05 considered significant in all analyses.

## RESULTS

From 2020-2022, we enrolled 140 participants; 122 had complete blood pressure data from TB treatment initiation and were included in our analyses. Study participants were mostly male (57%), <40 years (64%), and had DS/Hr TB (75%). Prevalence of hypertension at TB treatment intiation was 20% (24/122, 95% confidence interval [CI] 14–28). Among 98 participants without prevalent hypertension, 94 had ≥2 follow-up BP readings for incident hypertension classification. Of these, 26 developed hypertension during treatment or within 12 months of treatment completion (26/94, cumulative incidence=28%, 95%CI 20–37).

The unadjusted cumulative incidence of hypertension was significantly higher among participants ≥40 years (CIR=2.8, 95%CI 1.3–6.0), males (CIR=2.6, 95%CI 1.1– 7.1), and those with ≥1 cavitary lesion (CIR=2.4, 95%CI 1.1–5.1) or VAI ≥2.60 (CIR=2.2, 95%CI 1.2–7.7) (Table 1). Although non-significant, the cumulative incidence of hypertension among those with ≥1 cavitary lesion was 2.3 times (95%CI 0.9–6.2) the incidence among those without cavitary lesion after adjusting for age, sex, drug-susceptibilty profile, 5% relative BMI increase during TB treatment, and lipid profile. The CIR estimated with the WHO-defined hypertension cut-off was similar to the more sensitive cut-off used in our primary analyses (Table S2).

**Table 1.** Factors associated with incident hypertension among adults successfully treated for confirmed pulmonary tuberculosis in the country of Georgia, 2020–2022 (n=94)

| Characteristics | Total<br>N = 94 | Incidence<br>Hypertension |  | Cumulative Incidence Ratio |  |
| --- | --- | --- | --- | --- | --- |
|  |  | No<br>N (%) = 68 | Yes<br>N (%) = 26 | Crude CIR<br>(95%CI) | Adjusted<sup>a</sup><br>CIR (95%CI) |
| Age group<sup>b</sup> |  |  |  |  |  |
| <40 | 69 (73) | 56 (81) | 13 (19) | Ref | Ref |
| ≥40 | 25 (27) | 12 (48) | 13 (52) | **2.8 (1.3–6.0)** | 2.5 (0.9–6.5) |
| Sex<sup>b</sup> |  |  |  |  |  |
| Female | 41 (44) | 35 (85) | 6 (15) | Ref | Ref |
| Male | 53 (56) | 33 (62) | 20 (38) | **2.6 (1.1–7.1)** | 2.0 (0.7–6.4) |
| DST Profile<sup>b</sup> |  |  |  |  |  |
| DSTB/Hr TB<sup>c</sup> | 70 (74) | 50 (71) | 20 (29) | Ref | Ref |
| RR/MDR TB+<sup>d</sup> | 24 (26) | 18 (75) | 6 (25) | 0.9 (0.3–2.1) | 0.9 (0.3–2.7) |
| BMI<sup>b, e</sup> |  |  |  |  |  |
| Underweight | 26 (28) | 23 (88) | 3 (12) | 0.4 (0.1–1.2) |  |
| Normal | 53 (56) | 37 (70) | 16 (30) | Ref |  |
| Overweight/Obese | 9 (10) | 5 (56) | 4 (44) | 1.5 (0.4–4.0) |  |
| Missing | 6 (6) | 3 (50) | 3 (50) |  |  |
| BMI increase by ≥5%<br>during treatment<sup>b</sup> |  |  |  |  |  |
| No | 45 (48) | 32 (71) | 13 (29) | Ref | Ref |
| Yes | 43 (46) | 33 (77) | 10 (23) | 0.8 (0.3–1.8) | 2.5 (0.5–8.6) |
| Missing | 6 (6) | 3 (5) | 3 (5) |  |  |
| Cavitary lesion(s)<sup>f</sup> |  |  |  |  |  |
| None | 69 (73) | 55 (80) | 14 (20) | Ref | Ref |
| ≥1 | 25 (27) | 13 (52) | 12 (48) | **2.4 (1.1–5.1)** | 2.3 (0.9–6.2) |
| Severe lobe(s)<sup>f, g</sup> |  |  |  |  |  |
| None | 81 (86) | 60 (74) | 21 (26) | Ref |  |
| ≥1 | 13 (14) | 8 (62) | 5 (38) | 1.5 (0.5–3.6) |  |
| Total lung severity<br>score<sup>f, h</sup> |  |  |  |  |  |
| <10 | 87 (93) | 63 (72) | 24 (28) | Ref |  |
| ≥10 | 7 (7) | 5 (71) | 2 (29) | 1.0 (0.9–1.1) |  |
| Elevated LDL<sup>f, i</sup> (n=87) |  |  |  |  |  |
| No | 83 (88) | 63 (76) | 20 (24) | Ref | Ref |
| Yes | 4 (4) | 1 (25) | 3 (75) | 3.1 (0.7–9.1) | 2.5 (0.5–8.6) |
| Missing | 7 (7) | 4 (6) | 3 (4) |  |  |
| Low HDL<sup>f, j</sup> (n=87) |  |  |  |  |  |
| No | 45 (48) | 36 (80) | 9 (20) | Ref | Ref |
| Yes | 42 (45) | 28 (67) | 14 (33) | 1.7 (0.7–4.0) | 1.0 (0.4–3.0) |
| Missing | 7 (7) | 4 (6) | 3 (4) |  |  |
| Elevated TGL<sup>f, k</sup> |  |  |  |  |  |
| No | 68 (82) | 59 (77) | 18 (23) | Ref |  |
| Yes | 26 (18) | 9 (53) | 8 (47) | 2.0 (0.8–4.5) |  |
| Elevated HbA1c<sup>b, l</sup> |  |  |  |  |  |
| No | 86 (91) | 62 (72) | 24 (28) | Ref |  |
| Yes | 8 (9) | 6 (75) | 2 (25) | 0.9 (0.1–3.0) |  |
| Visceral Adipose Index<sup>b, m</sup> |  |  |  |  |  |
| ≤2.60 | 43 (46) | 37 (86) | 6 (14) | **Ref** |  |
| >2.60 | 44 (47) | 27 (61) | 17 (39) | **2.8 (1.2–7.7)** |  |
| Missing | 7 (7) | 4 (6) | 3 (4) |  |  |

<sup>a</sup> Results from fully-adjusted multivariable model (all predictors were included in the final model)
<sup>b</sup> Collected at TB treatment initiation
<sup>c</sup> Included individuals with drug-susceptible and isoniazid mono-resistant TB (DS/Hr TB)
<sup>d</sup> Included individuals with rifampicin-resistant and multidrug/extensively drug-resistant TB (RR/MDR TB+)
<sup>e</sup> Underweight: <18.5 kg/m<sup>2</sup>; Normal: 18.5–24.9 kg/m<sup>2</sup>; Overweight/obese: ≥25 kg/m<sup>2</sup>
<sup>f</sup> Collected at TB treatment completion
<sup>g</sup> Had at least one lung lobe with >75% involvement
<sup>h</sup> Composite impairment score of 5 pulmonary lobes (lung involvement 0–25%: 1, 26–50%: 2, 51–75%: 3, 76–100%: 4, maximum score=20) was classified into mild (score 1–10) or moderate/severe (score >10).
<sup>i</sup> ≥130 mg/dL
<sup>j</sup> <40 mg/dL for men and <50 mg/dL for women
<sup>k</sup> ≥150 mg/dL
<sup>l</sup> ≥5.7%
<sup>m</sup> Cut-point based on median value
CIR – cumulative incidence ratio; DST – drug-susceptibility testing; DSTB – drug-susceptible tuberculosis; Hr– isoniazid-resistant; MDR – multidrug resistant tuberculosis; TB – tuberculosis; BMI – body mass index; LDL – low-density lipoprotein; HDL – high-density lipoprotein; TGL – triglycerides; HbA1c – hemoglobin A1c

Among participants included in the incidence analyses and available SBP levels at TB treatment initiation (n=91), the median change in SBP was 10mmHg (IQR 0–19), 5mmHg (IQR −2.0–10.0mmHg) among participants without incident hypertension, and 20mmHg (IQR 10.2–30.0mmHg) among participants with incident hypertesion. Among those with incident hypertension, the median change in SBP was slightly higher among those with ≥1 cavitary lesions (Figure 1A) or ≥1 severe lobe involvement (Figure 1B) compared to those without.

**Figure 1.**
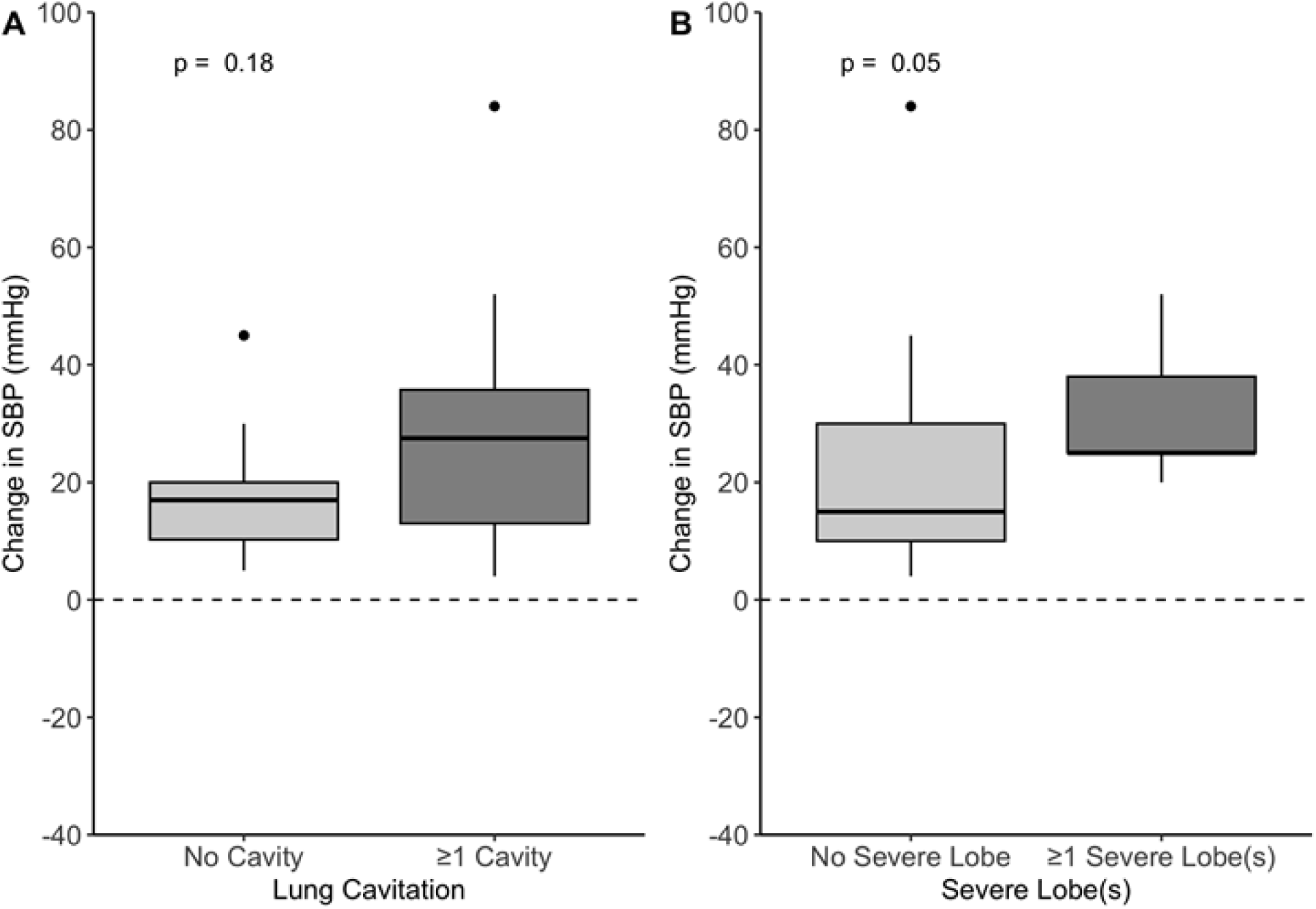
Median change in systolic blood pressure (SBP) from tuberculosis treatment initiation to 12-month post-tuberculosis treatment completion among individuals with incident hypertension, Tbilisi, Georgia 2020–2022 (N=26) Boxes represent the interquartile range (IQR), whiskers represent data falling within 1.5 times IQR, and points represent outliers (>1.5 times IQR). Panel A) presence of cavitary lesions (No cavity median: 17.0mmHg vs. ≥1 cavity median: 27.5mmHg, median difference=10.5mmHg), and Panel B) presence of severe lobe(s) (No severe lobe median: 15.0mmHg vs. ≥1 severe lobe(s) median: 25.0mmHg, median difference=10.0mmHg).

## DISCUSSION

In our cohort, one-fifth had hypertension at TB treatment initiation, and more than one-quarter of participants without prevalent hypertension developed incident hypertension during or within 12 months of completing treatment. Our study is the first to assess the association between cavitary lesions and hypertension incidence, highlighting the need for larger prospective cohort studies to comprehensively assess the impact of TB-severity factors on hypertension or CVD risks post-TB treatment.

Our study suggested that the cumulative incidence risk of hypertension among participants with ≥1 cavitary lesion may be twice the risk among participants without cavitary lesions (adjusted CIR was non-significant). More severe TB manifestations (e.g., marked by lung cavitation) may lead to a prolonged or greater degree of systematic inflammation, a key factor in the development of hypertension.^16,17^ Individuals with severe manifestations of TB may benefit from routine screening for hypertension to monitor of cardiovascular risk trajectories while in TB care.

Our preliminary findings highlight an important relationship between TB and hypertension. Although our study sample was small and from a single site, our findings suggest that it is critical to improve our understanding of the hypertension burden and trajectories of CVD risks post-TB treatment. Routinely measuring blood pressure levels during and at the end of TB treatment could be an inexpensive way to identify individuals with early signs of CVD and determine whether referrals to other clinics are needed. Future studies should aim to measure how integrating post-TB treatment care and NCD programs (e.g., NCD screening, referral to NCD clinics) impacts post-TB morbidity and mortality.

## DECLARATIONS AND ACKNOWLEGEMENTS

## Acknowledgments

The authors thank the study doctors (Leila Goginashvili, Sergo Vashakidze, Nino Jakobia, Manana Rekhviashvili, Khatuna Guchmazashvili, Marine Gachechiladze, Liana Tsivtsivadze, Tamar Natriashvili) who helped the participant enrollment process at the National Center for Tuberculosis and Lung Diseases, Tbilisi, Georgia.

## Potential Conflicts of Interest

We have no conflict of interest to declare.

## Patient Consent Statement

The study was submitted to, reviewed, and approved by the Institutional Review Boards (IRBs) at Emory University, Atlanta, USA, and the ethics committee at the National Center for Tuberculosis and Lung Diseases, Tbilisi, Georgia (FWA000020831). All participants provided written informed consent prior to study participation.

## Funding

This work was supported in part by grants from the National Institutes of Health (NIH) including the National Institute of Allergy and Infectious Diseases (NIAID) [R03AI133172 to MJM, R03AI139871 to RRK, R01AI153152 to MJM], the National Heart, Lung, and Blood Institute (NHLBI) [R01HL156779 to MAH], and the Fogarty International Center (FIC) [R21TW011157 to MK and MJM]. ADS was supported by a Vanderbilt Emory Cornell Duke (VECD) Global Health Fellowship, funded by the NIH FIC (D43TW009337). The content is solely the responsibility of the authors and does not necessarily represent the official views of the National Institutes of Health.

## Authors Contributions

MJM, ADS and VCO conceived the study design. MK, TA, MG and ADS collected the data. VCO performed the analyses. VCO, MJM, and ADS interpreted the results. VCO and ADS wrote the first draft of the manuscript. MJM assisted with further drafting and revisions of manuscripts. All authors reviewed and approved the final version of the manuscript.

## Data Availability Statement

The data that support the findings of this study are available from the corresponding author, ADS, upon reasonable request.

## SUPPLEMENTAL MATERIALS

**Table S1.** Comparison of Akaike Information Criterion (AIC) values across multivariable models.

| **Models** | **Covariates included** | **Cavitary Lesion(s)** | **aCIR (95%CI)** | **AIC** |
| --- | --- | --- | --- | --- |
| Model 1<sup>a</sup> | age group, gender, DST profile, BMI increase by $\geq 5\%$ during treatment, LDL, HDL | None $\geq 1$ | Reference<br>2.3 (0.9–6.2) | 99.2 |
| Model 2 | age group, gender, DST profile, BMI increase by $\geq 5\%$ during treatment, LDL, HDL, total lung severity score | None $\geq 1$ | Reference<br>2.8 ( 0.9–7.9) | 100.6 |
| Model 3 | age group, gender, DST profile, BMI increase by $\geq 5\%$ during treatment, LDL, HDL, severe lobe(s) | None $\geq 1$ | Reference<br>2.2 (0.7–6.3) | 101.06 |
| Model 4 | age group, gender, DST profile, VAI | None $\geq 1$ | Reference<br>2.3 (1.0–5.4) | 102.4 |
| Model 5 | age group, gender, DST profile, BMI, LDL, HDL | None $\geq 1$ | Reference<br>2.6 (1.1–6.1) | 107.2 |
| <sup>a</sup> Reported as final model in text |  |  |  |  |

**Table S2.**
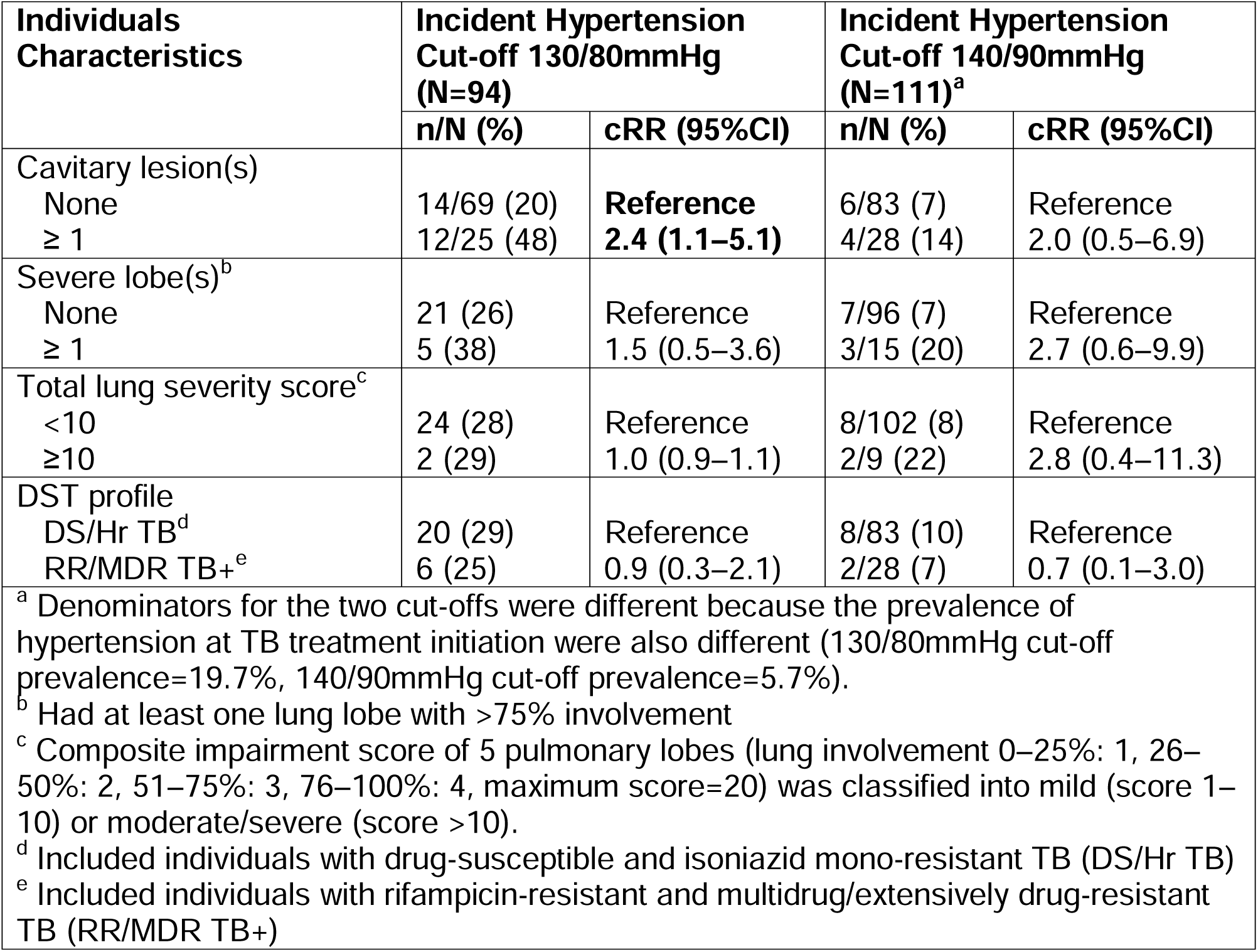
Factors associated with incident hypertension (BP≥140/90) among adults successfully treated for confirmed pulmonary tuberculosis in the country of Georgia, 2020–2022.

## Notes

### Competing Interest Statement

The authors have declared no competing interest.

### Author Declarations

The study was submitted to, reviewed, and approved by the Institutional Review Boards (IRBs) at Emory University, Atlanta, USA, and the ethics committee at the National Center for Tuberculosis and Lung Diseases, Tbilisi, Georgia (FWA000020831).

